# DESCRIPTION OF THE METABOLIC SYNDROME AND ITS INDIVIDUALIZED RISK IN A LATIN AMERICAN COHORT OF GERIATRIC PATIENTS WITH HYPOTHYROIDISM

**DOI:** 10.1101/2023.08.31.23294482

**Authors:** Luis Andres Dulcey Sarmiento, Juan Sebastián Theran Leon, Jaime Gomez, Rafael Guillermo Parales Strauch, Raimondo Caltagirone, Edgar Camilo Blanco Pimiento, María Paula Ciliberti Artavia, Juan Camilo Martinez, Valentina Cabrera Peña, Maria Camila Amaya, Jorge Luis Vargas, Angie Lizcano, Aldahir Quintero, Juan Camilo Mayorca, Carlos Hernandez, Maria Alejandra Cala

## Abstract

**Introduction:** It is still discussed whether hypothyroidism as an independent risk factor for cardiovascular events. Our objective was to evaluate the association between hypothyroidism and the risks of cardiovascular events and mortality through 3 stratification systems.

**Methods:** A retrospective study was carried out by reviewing medical records in the period of January 2015 - December 2017 in a South American hospital. Patients had fasting total cholesterol, high-density lipoprotein (HDL) cholesterol, triglycerides and plasma glucose were included. Quantitative variables are presented as mean ± standard deviation or median (interquartile range) according to their distribution, and qualitative variables as percentages. Student’s test was performed to assess the differences between two variables. all statistical analyzes of the database results were performed with (SPSS for Windows, v.20.1; Chicago, IL).

**Results:** The present study has demonstrated metabolic syndrome criteria present in patients diagnosed with hypothyroidism in a high proportion. The male gender was 32% compared to the female 68%. The Framingham equation classified a higher percentage of women patients with hypothyroidism as low cardiovascular risk compared to the PROCAM and SCORE equations. Item was found that there was a greater cardiovascular risk in those patients with still uncontrolled hypothyroidism profile, showing a statistical correlation of this alteration for the 3 stratification systems used.

**Conclusions:** Hypothyroidism is a risk factor for cardiovascular disease. Uncontrolled hypothyroidism in the present study is associated with greater adverse outcomes in the medium and long term, the present study warns about the need to better characterize this patient cohorts.

## Introduction

Thyroid hormones have extensive effects on the body and play an important role in the homeostasis of the cardiovascular system (1). Abnormal thyroid function has consequences for the health of the general public. Patients with hypothyroidism are at increased risk of cardiovascular abnormalities, such as dyslipidemia and accelerated atherosclerosis (2).

It is not fully known whether hypothyroidism is an independent risk factor for cardiovascular events or mortality, since there are conflicting opinions in this regard. It is also contradictory whether adults with hypothyroidism and pre-existing cardiovascular disease might be at particularly high cardiovascular risk (3-5). Although there have been meta-analyses and even meta-analyses of individual participant data on the subject so far (6-10), they have been restricted to small cohorts. Therefore, the present study is justified, which seeks to establish, through three stratification systems, in a comparative way, cardiovascular risk and the presence of metabolic syndrome alterations in a cohort of South American patients.

## Materials and methods

### Population

The hypothyroidism group consisted of a random sample of 100 consecutive recruited patients. The diagnosis of hypothyroidism was confirmed according to the criteria published by the American Thyroid Association (11) and the hormonal diagnosis was based on a detection kit through ELISA with a thyroid panel (free T4 and TSH) from the commercial house MONOBIND INC which has been validated internationally (12). Patients older than 65 years were included since the cardiovascular risk equations used have not been validated in younger patients.

### Methods

was carried out by reviewing medical records in the period of January 2015 - December 2017 in a South American hospital. Patients who had fasting total cholesterol, high-density lipoprotein (HDL) cholesterol, triglycerides, and plasma glucose were included.

### Cardiovascular risk assessment

The frequency and factors of syndrome metabolism were analyzed using the ATP-III criteria. Cardiovascular risk was estimated for each subject using the three risk equations and subjects were then classified as having low, moderate, or high 10-year coronary risk using Framingham, PROCAM (<10%, 10%-20%, and > 20 %, respectively) and SCORE (<3%, 3% -4% and ≥5%, respectively) (13). Patients with established coronary artery disease or other atherosclerotic disease were directly defined as having high cardiovascular risk (>20%) according to all three guidelines (14). The same occurred with those in primary prevention with type 2 diabetes mellitus, when the Framingham and SCORE systems were used (15). On the other hand, the risk SCORE was calculated in patients infected with and without hypothyroidism with risk factors from 0 to 1 in primary prevention, in whom risk assessment was not necessary when the NCEP-ATP III guidelines were applied (15). For the SCORE system, the 10-year graph calculated the risk of cardiovascular disease by sex, age, systolic blood pressure, smoking status, and total cholesterol/HDL cholesterol ratio (15).

### Statistic analysis

Quantitative variables are presented as mean ± standard deviation or median (interquartile range) according to their distribution, and qualitative variables as percentages with 95% confidence intervals (CI). Student ‘s t test was performed to assess the differences between two variables. A value of p < 0.05 was considered statistically significant. All statistical analyzes of the database results were performed using the Statistical Package for the Social Sciences (SPSS for Windows, v.20.1; Chicago, IL).

### Ethical Considerations

The ethical aspects of this research work were carried out based on the criteria of the *Belmont Report*, adjusted to its principles of respect for the person, beneficence and justice, and the Declaration of Helsinki of the World Medical Association of 1964. Likewise, will be subject to Venezuelan legislation and its 1985 code of medical ethics (currently in force) in its title V, chapter 4, regarding research in human beings. The highest standards were maintained that allowed the protection of the privacy and physical integrity of the participants. Through informed consent, the objectives of this research were explained, in the same way, the procedures that were performed and the inherent risks and complications were explained in detail and in clear and understandable language. The ethics committee of the postgraduate course in Internal Medicine of the Universidad de los Andes in meeting 009-2018 endorsed the carrying out of the study since it had all the ethical aspects in the methodological approaches, once it was completed the go-ahead was given for the publication of the study.

## Results

The baseline characteristics of the population included in the study are presented, as well as the time of diagnosis and the mean dose of levothyroxine (Table 1).

| Variable | Values |
| --- | --- |
| Ages | 79.5 ± 14.1 years |
| Gender | Female 68% Male 32% |
| Diagnosis time of hypothyroidism | 11.8 ± 5.6 years |
| Mean dose of levothyroxine | 75 ± 51 mcg |

Regarding the ATP-III criteria, these were the findings found in the cohort of 100 patients (Table 2).

**Table 2.**
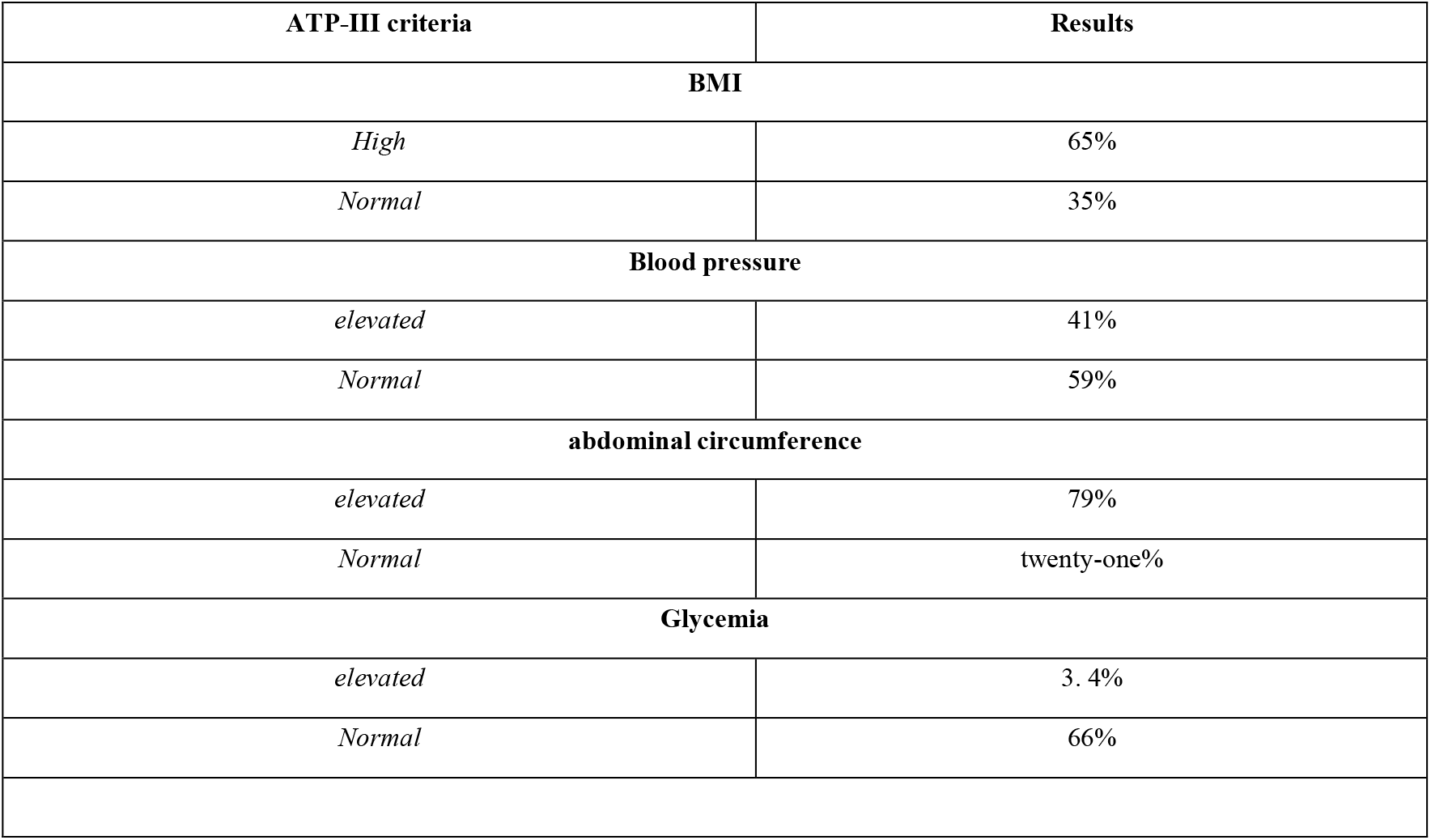
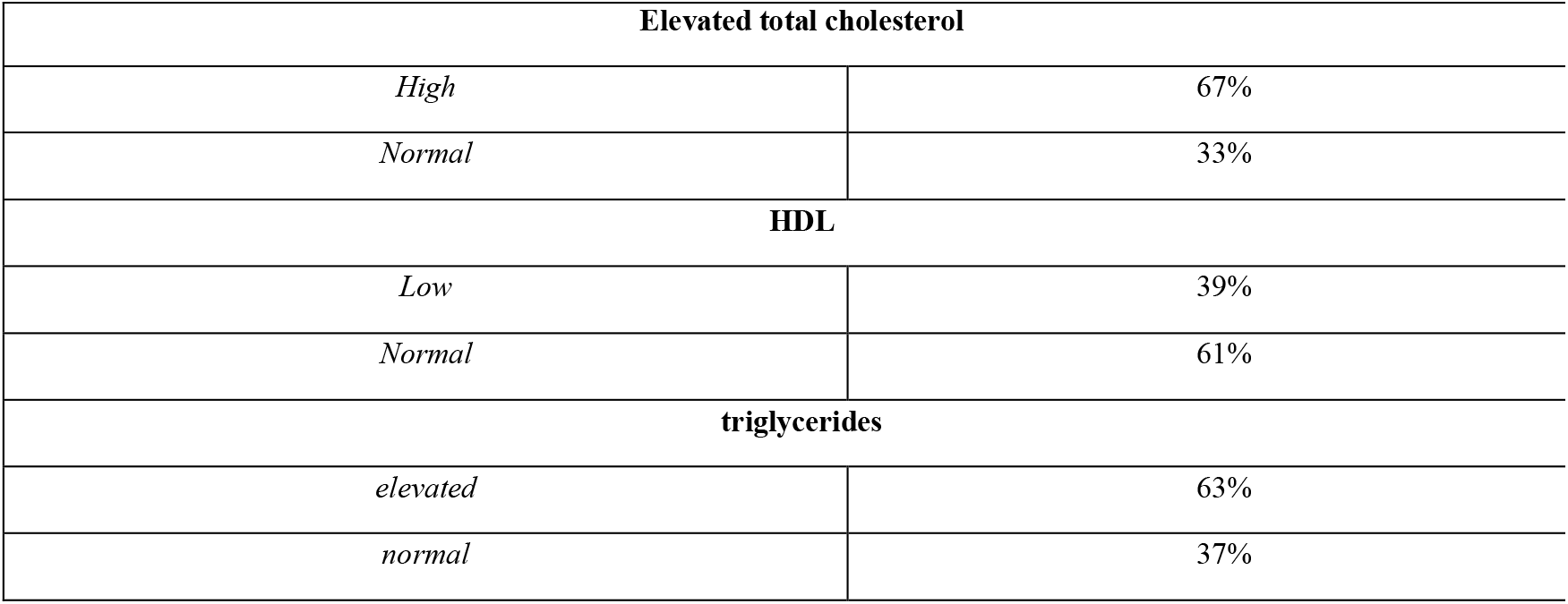
ATP-III criteria and results of the patient cohort.

Table **3** shows the cardiovascular risk estimated by the 3 scores in terms of low, moderate and high risk modalities.

**Table 3.** Prevalence of patients with hypothyroidism with low, moderate and high cardiovascular risk according to Framingham (<10%, 10% -20% and >20%), SCORE (<3%, 3% -4% and ≥5%). and PROCAM (<10%, 10%-20% and >20%).

| Risk | Low | Moderate | High |
| --- | --- | --- | --- |
| **FRAMINGHAM** | 71% | fifteen% | 14% |
| **SCORE** | 64% | 22% | 14% |
| **PROCAM** | 59% | 28% | 13% |

In relation to the values of cardiovascular risk by gender, these variables are presented in **(Table 4)**.

**Table 4.** Prevalence of patients classified as low, moderate, and high cardiovascular risk according to gender according to FRAMINGHAM (<10%, 10% -20%, and >20%, respectively), SCORE (<3%, 3% -4%, and ≥5 %, respectively), PROCAM (<10%, 10% – 20% and >20%, respectively)

| Scores and risk | Women | Men |
| --- | --- | --- |
| Framingham |  |  |
| **Low** | 52% | 19% |
| **Moderate** | 10% | 5% |
| **High** | 6% | 8% |

| SCORE |  |  |
| --- | --- | --- |
| Low | 48% | 16% |
| Moderate | fifteen% | 7% |
| High | 5% | 9% |
| PROCAM |  |  |
| Low | 53% | 6% |
| Moderate | eleven% | 17% |
| High | 4% | 9% |

Table **5** shows the correlation between control of the thyroid profile and cardiovascular risk according to the 3 scores.

**Table 5.** Correlation between control of hypothyroidism and cardiovascular risk according to the 3 scores.

| variables | p value for Low risk |  | p value for Moderate risk |  | p value for High risk |  |
| --- | --- | --- | --- | --- | --- | --- |
| **Framingham** | controlled<br>hypothyroidism<br>p = 0.089 | Uncontrolled<br>hypothyroidism<br>p = 0.075 | controlled<br>hypothyroidism<br>p = 0.154 | Uncontrolled<br>hypothyroidism<br>p = 0.128 | controlled<br>hypothyroidism<br>p = 0.052 | Uncontrolled<br>hypothyroidism<br>**p = 0.046 |
| **SCORE** | controlled<br>hypothyroidism<br>p = 0.102 | Uncontrolled<br>hypothyroidism<br>p = 0.119 | controlled<br>hypothyroidism<br>p = 0.095 | Uncontrolled<br>hypothyroidism<br>p = 0.101 | controlled<br>hypothyroidism<br>p = 0.061 | Uncontrolled<br>hypothyroidism<br>**p = 0.043 |
| **PROCAM** | controlled<br>hypothyroidism<br>p = 0.115 | Uncontrolled<br>hypothyroidism<br>p = 0.131 | controlled<br>hypothyroidism<br>p = 0.087 | Uncontrolled<br>hypothyroidism<br>p = 0.099 | controlled<br>hypothyroidism<br>p = 0.055 | Uncontrolled<br>hypothyroidism<br>**p = 0.039 |

**\*\*Statistical significance found in the analysis between variables.**

## Discussion

The present study demonstrated that the presence of metabolic syndrome criteria in patients diagnosed with hypothyroidism is really high. The female gender was 68% compared to the female 32%. The time of diagnosis of hypothyroidism was 11.8 + 5.6 years. The presence of abdominal circumference was the main criterion of metabolic syndrome, followed by high total cholesterol and in third place, high body mass index levels; these metabolic alterations have been found in the population with hypothyroidism internationally (16). These values are worrisome since they indicate a high prevalence of metabolic syndrome criteria in this particular population cohort.

Framingham equation classified a higher percentage of female patients with hypothyroidism as low cardiovascular risk compared to the PROCAM and SCORE equations. This study showed a high prevalence of hypothyroidism patients with low cardiovascular risk regardless of the cardiovascular risk system used.

The concordance observed between Framingham and PROCAM was 84% (w 0.45, p <0.0001), between Framingham and SCORE 79.2% (w 0.37, p <0.0001) and between PROCAM and SCORE 90.4 % (p 0.42, p < 0.0001). On the other hand, leaving aside the comparisons of risk equations, the different scales for estimating global cardiovascular risk in the general population could underestimate the real risk in patients with hypothyroidism, some studies point out (17). The values of high cardiovascular risk estimated by gender were almost comparable for the 3 stratification systems used. The estimate of cardiovascular risk in this particular group of patients may be underestimated. It was found that there was a greater cardiovascular risk in those patients with uncontrolled hypothyroidism, showing statistical correlation. These findings are consistent with the higher mortality, including cardiovascular mortality, in those who do not present control of the disease, as has been demonstrated in other studies (18-20). The present study emphasizes the need to validate the different risk equations used to assess cardiovascular risk in patients with hypothyroidism and to adapt the proposal to include thyroid profile levels in the comprehensive evaluation.

## Conclusions

Global risk assessment has become an accepted component of clinical guidelines and recommendations in cardiovascular medicine. The metabolic syndrome conditions a greater cardiovascular risk in the general population, this bidirectional relationship does not escape the scope of those with hypothyroidism. Especially uncontrolled hypothyroidism shows an association with increased risk of coronary disease across all 3 stratification systems used in the present study. Adequate replacement with hormone replacement therapy in patients with hypothyroidism is a fundamental strategy to reduce potential risks.

This study is limited by multiple variables. In the first place because of its retrospective and cross-sectional design in a single center. Second, we do not prospectively follow up on clinical events in patients. Third, we do not follow patients for metabolic control over time. Fourth, some of our analyzes may be limited by a small sample size as well as the small volume in the subgroup of men who have a higher cardiovascular risk at younger ages, which can lead to bias when doing the analyses. We considered carrying out studies with larger samples of patients and reviewing the scientific evidence derived a posteriori in the characterization of this population cohort.

## Data Availability

All data produced in the present work are contained in the manuscript

## Notes

### Competing Interest Statement

The authors have declared no competing interest.

### Funding Statement

This study did not receive any funding

### Author Declarations

The ethics committee of the postgraduate course in Internal Medicine of the Universidad de los Andes in meeting 009-2018 endorsed the carrying out of the study since it had all the ethical aspects in the methodological approaches, once it was completed the go-ahead was given for the publication of the study.

